# Childhood adversity is associated with longitudinal white matter changes after adulthood trauma

**DOI:** 10.1101/2025.03.08.25323425

**Authors:** Tianyi Li, Megan E. Huibregtse, Timothy D. Ely, Sanne J H. van Rooij, Lauren A M. Lebois, E. Kate Webb, Tanja Jovanovic, Stacey L. House, Steven E. Bruce, Vishnu P. Murty, Francesca L. Beaudoin, Xinming An, Thomas C. Neylan, Gari D. Clifford, Sarah D. Linnstaedt, Kenneth A. Bollen, Scott L. Rauch, John P. Haran, Alan B. Storrow, Christopher Lewandowski, Paul I. Musey, Phyllis L. Hendry, Sophia Sheikh, Christopher W. Jones, Brittany E. Punches, Lauren A. Hudak, Jose L. Pascual, Mark J. Seamon, Elizabeth M. Datner, Claire Pearson, David A. Peak, Roland C. Merchant, Robert M. Domeier, Niels K. Rathlev, Brian J. O’Neil, Paulina Sergot, Leon D. Sanchez, John F. Sheridan, Ronald C. Kessler, Karestan C. Koenen, Kerry J. Ressler, Samuel A. McLean, Jennifer S. Stevens, Nathaniel G. Harnett

**Affiliations:** Division of Depression and Anxiety, McLean Hospital, Belmont, MA, 02478, USA; Department of Psychiatry and Behavioral Sciences, Emory University School of Medicine, Atlanta, GA, 30329, USA; Department of Psychiatry, Harvard Medical School, Boston, MA, 02115, USA; Division of Depression and Anxiety Disorders, McLean Hospital, Belmont, MA, 02478, USA; Department of Psychiatry and Behavioral Neurosciences, Wayne State University, Detroit, MI, 48202, USA; Department of Emergency Medicine, Washington University School of Medicine, St. Louis, MO, 63110, USA; Department of Psychological Sciences, University of Missouri - St. Louis, St. Louis, MO, 63121, USA; Department of Psychology, Temple University, Philadelphia, PA, 19121, USA; Department of Epidemiology, Brown University, Providence, RI, 02930, USA; Department of Emergency Medicine, Brown University, Providence, RI, 02930, USA; Institute for Trauma Recovery, University of North Carolina at Chapel Hill, Chapel Hill, NC, 27559, USA; Department of Anesthesiology, University of North Carolina at Chapel Hill, Chapel Hill, NC, 27559, USA; Departments of Psychiatry and Neurology, University of California San Francisco, San Francisco, CA, 94143, USA; Department of Biomedical Informatics, Emory University School of Medicine, Atlanta, GA, 30332, USA; Department of Biomedical Engineering, Georgia Institute of Technology and Emory University, Atlanta, GA, 30332, USA; Department of Psychology and Neuroscience & Department of Sociology, University of North Carolina at Chapel Hill, Chapel Hill, NC, 27559, USA; Institute for Technology in Psychiatry, McLean Hospital, Belmont, MA, 02478, USA; Department of Psychiatry, McLean Hospital, Belmont, MA, 02478, USA; Department of Emergency Medicine, University of Massachusetts Chan Medical School, Worcester, MA, 01655, USA; Department of Emergency Medicine, Vanderbilt University Medical Center, Nashville, TN, 37232, USA; Department of Emergency Medicine, Henry Ford Health System, Detroit, MI, 48202, USA; Department of Emergency Medicine, Indiana University School of Medicine, Indianapolis, IN, 46202, USA; Department of Emergency Medicine, University of Florida College of Medicine -Jacksonville, Jacksonville, FL, 32209, USA; Department of Emergency Medicine, Cooper Medical School of Rowan University, Camden, NJ, 08103, USA; Department of Emergency Medicine, Ohio State University College of Medicine, Columbus, OH, 43210, USA; Ohio State University College of Nursing, Columbus, OH, 43210, USA; Department of Emergency Medicine, Emory University School of Medicine, Atlanta, GA, 30329, USA; Department of Surgery, Department of Neurosurgery, University of Pennsylvania, Philadelphia, PA, 19104, USA; Perelman School of Medicine, University of Pennsylvania, Philadelphia, PA, 19104, USA; Department of Surgery, Division of Traumatology, Surgical Critical Care and Emergency Surgery, University of Pennsylvania, Philadelphia, PA, 19104, USA; Department of Emergency Medicine, Jefferson Einstein hospital, Jefferson Health, Philadelphia, PA, 19141, USA; Department of Emergency Medicine, Sidney Kimmel Medical College, Thomas Jefferson University, Philadelphia, PA, 19107, USA; Department of Emergency Medicine, Wayne State University, Ascension St. John Hospital, Detroit, MI, 48236, USA; Department of Emergency Medicine, Massachusetts General Hospital, Boston, MA, 02114, USA; Department of Emergency Medicine, Harvard Medical School, Boston, MA, 02115, USA; Department of Emergency Medicine, Brigham and Women’s Hospital, Boston, MA, 02115, USA; Department of Emergency Medicine, Trinity Health-Ann Arbor, Ypsilanti, MI, 48197, USA; Department of Emergency Medicine, University of Massachusetts Medical School-Baystate, Springfield, MA, 01107, USA; Department of Emergency Medicine, Wayne State University, Detroit Receiving Hospital, Detroit, MI, 48202, USA; Department of Emergency Medicine, McGovern Medical School at UTHealth, Houston, TX, 77030, USA; Division of Biosciences, Ohio State University College of Dentistry, Columbus, OH, 43210, USA; Institute for Behavioral Medicine Research, OSU Wexner Medical Center, Columbus, OH, 43211, USA; Department of Health Care Policy, Harvard Medical School, Boston, MA, 02115, USA; Department of Epidemiology, Harvard T.H. Chan School of Public Health, Harvard University, Boston, MA, 02115, USA; Department of Emergency Medicine, University of North Carolina at Chapel Hill, Chapel Hill, NC, 27559, USA; Department of Psychiatry, University of North Carolina at Chapel Hill, Chapel Hill, NC, 27559, USA

**Keywords:** Childhood Adversity, White Matter, PTSD, Diffusion Tensor Imaging

## Abstract

**Background:** Childhood adversity is associated with susceptibility to posttraumatic stress disorder (PTSD) in adulthood. Both PTSD and adverse experiences in childhood are linked to disrupted white matter microstructure, yet the role of white matter as a potential neural mechanism connecting childhood adversity to PTSD remains unclear. The present study investigated the potential moderating role of previous childhood adversity on longitudinal changes in white matter microstructures and posttraumatic stress symptoms following a recent traumatic event in adulthood.

**Methods:** As part of the AURORA Study, 114 recent trauma survivors completed diffusion weighted imaging at 2-weeks and 6-months after exposure. Participants reported on prior childhood adversity and PTSD symptoms at 2-weeks, 6-months, and 12-months post-trauma. We performed both region-of-interest (ROI) and whole-brain correlational tractography analyses to index associations between white matter microstructure changes and prior adversity.

**Results:** Whole-brain correlational tractography revealed that greater childhood adversity moderated the changes in quantitative anisotropy (QA) over time across threat and visual processing tracts including the cingulum bundle and inferior fronto-occipital fasciculus (IFOF). Further, QA changes within cingulum bundle, IFOF, and inferior longitudinal fasciculus were associated with changes in PTSD symptoms between 2-weeks and 6-months.

**Conclusions:** Our findings suggest temporal variability in threat and visual white matter tracts may be a potential neural pathway through which childhood adversity confers risk to PTSD symptoms after adulthood trauma. Future studies should take the temporal properties of white matter into consideration to better understand the neurobiology of childhood adversity and PTSD.

## Introduction

Childhood adversity can potentiate later development of psychiatric disorders in adulthood. For example, experiences of childhood adversity are associated with higher risk of an adult-onset posttraumatic stress disorder (PTSD) diagnosis (McLaughlin et al., 2017), more severe PTSD symptoms in adulthood (Wong et al., 2023), and lower likelihood to recover from PTSD once diagnosed (Steinert et al., 2015). However, the potential neural mechanisms underlying the impact of childhood experiences on adulthood trauma responses are not well explored. Prior work suggests adverse experiences in childhood are tied to alterations in white matter development within threat neurocircuitry (Olson et al., 2020; Puetz et al., 2017; McLaughlin et el., 2019). Further, recent work suggests childhood trauma may alter the peritraumatic integrity of sensory-related white matter tracts which in turn influence later PTSD symptoms after an adulthood trauma (Wong et al., 2023). However, there remains little research on potential longitudinal changes in white matter microstructure after adulthood trauma (e.g., between the peri-and post-traumatic period) that may be influenced by childhood experiences. Childhood experiences may modulate the degree of white matter changes following later trauma and subsequently elevate neural risk for PTSD. Therefore, the present research investigated structural changes in threat and sensory white matter tracts in the early aftermath of adulthood trauma to better understand the influence of childhood adversity on brain structure that supports adulthood PTSD.

Both PTSD and childhood adversity are associated with structural alterations in threat and sensory neurocircuitry (Daniels et al., 2013). The core threat neurocircuit includes the prefrontal cortex, hippocampus, and amygdala, and is linked to the emergence and maintenance of PTSD symptoms (Harnett et al., 2020b; Roeckner et al., 2021). For example, previous investigations have observed reduced integrity of the cingulum bundle and uncinate fasciculus - white matter tracts that interlink threat-related regions - in individuals with PTSD (Fani et al., 2016; Olson et al., 2017; Koch et al., 2017). Childhood adversity is also associated with lesser microstructure of similar tracts such as the uncinate fasciculus, cingulum bundle, and corpus callosum in children, adolescents, and adults (Buimer et al., 2022; Kulla et al., 2024; Hanson et al., 2015; Huang et al., 2012). The prior work indicates structural pathways that support threat processing are important for understanding the neural impact of childhood adversity on later PTSD development. Recent research has also emphasized that sensorial circuitry – particularly visual circuitry – is also strongly related to both PTSD and childhood adversity (Harnett et al., 2024). Meta-analyses suggest individuals with PTSD show reduced fractional anisotropy (FA) of the superior longitudinal fasciculus and inferior fronto-occipital fasciculus (IFOF) compared with trauma-exposed controls (Siehl et al., 2018; Ju et al., 2020). In addition, more adverse experiences in childhood are associated with lower integrity of visual association tracts such as the IFOF, inferior longitudinal fasciculus (ILF), and superior longitudinal fasciculus (Tendolkar et al., 2017; Huang et al., 2012). The IFOF and ILF in particular are critical for interconnecting threat and visual circuitry along the ventral visual stream (Zemmoura et al., 2022; Forkel et al., 2014). Taken together, the current literature suggests that alterations in white matter fibers, especially those connecting threat and visual networks, may be a possible neural mechanism underlying the association between childhood adversity and PTSD.

Recent longitudinal investigations of recent trauma survivors further suggest childhood adversity is linked with neurobehavioral alterations across threat and sensorial circuits related to PTSD in the early aftermath of adulthood traumatic stress. A prior report found heightened amygdala-precuneus resting-state functional connectivity of amygdala 2-weeks post-trauma mediated the influence of childhood abuse on anxiety but not PTSD symptoms 6-months after trauma exposure (Harb et al., 2024). Similarly, previous research from our group observed that childhood maltreatment was associated with reduced integrity of the internal capsule following adulthood trauma which in turn predicted increased PTSD symptoms at 6-months post-trauma (Wong et al., 2023). These findings highlight the possibility that microstructural variability in the acute aftermath of trauma exposure is a potential mediator between childhood adverse experiences and posttraumatic outcomes. However, no prior study has investigated potential longitudinal changes in white matter in recent trauma survivors which may be linked to recovery from, or maintenance of, PTSD symptoms over time. Prior work by Kennis et al. (2015) found that patients with persistent PTSD showed increased FA in the dorsal cingulum but not those who recovered after treatment or the combat-exposed controls. Another longitudinal study observed heightened FA in the posterior cingulum bundle over time in individuals whose PTSD symptoms persisted (Zhang et al., 2012). These data suggest that changes in white matter microstructure are related to changes in PTSD symptomatology. Determining if longitudinal changes in white matter microstructure are associated with childhood adversity and future PTSD symptoms would improve our understanding of the neurobiological risk factors of PTSD.

Therefore, the current study investigated the interrelations between childhood adversity, microstructural changes in white matter acutely after adulthood trauma, and later PTSD symptoms in recent trauma survivors. We hypothesized that childhood adversity would moderate the microstructural changes within key threat (e.g., uncinate fasciculus, cingulum bundle) and visual (IFOF, ILF) white matter tracts between 2-weeks and 6-months post-trauma such that trauma survivors with higher level of childhood adversity would show greater changes in these tracts. We also hypothesized that greater microstructural changes of such white matter would be associated with greater increases in PTSD symptoms over the same period. The present findings provided insights into the potential long-term effects of early adversity on the progression of PTSD symptoms after adulthood trauma and the underlying role of white matter integrity.

## Methods and Materials

### Participants

The present analysis utilized data from the Advancing Understanding of RecOvery afteR trauma (AURORA) Study, a longitudinal multisite investigation of neuropsychiatric outcomes in recent trauma survivors. Details of the broader AURORA study are available in prior reports (McLean et al., 2020). Individuals from the dataset were included in prior studies (Harnett et al., 2022; Wong et al., 2023); however, the present analyses are distinct. Briefly, volunteers aged between 18-75 who experienced a recent trauma were enrolled from Emergency Departments (ED) across the United States. Qualifying trauma exposures included motor vehicle collision, fall greater than 10 feet, sexual assault, physical assault, and mass casualty incident. Participants were also recruited if a) they endorsed having other life-threatening traumatic events on a screener question and b) a research assistant confirmed the event was qualifying. Exclusion criteria involved use of general anesthesia, long bone fracture, laceration with severe hemorrhage, solid organ injury exceeding American Association for the Surgery of Trauma Grade 1, lack of alertness and orientation at enrollment, visual or auditory impairments that hindered the completion of web-based assessments and/or follow-ups via telephone, injuries resulting from self-harm or occupational incidents, incarceration, ongoing domestic violence reported, intake of more than 20 mg morphine or its equivalent per day, lack of IOS or Android-compatible smartphone with Internet access, and lack of email address that could be checked as needed. Additional MRI exclusion criteria included having metal or ferromagnetic implants, current pregnancy or breastfeeding, claustrophobia, and a history of neurological disorders such as seizure, epilepsy, Parkinson’s disease, dementia, and Alzheimer’s disease. Written informed consent was obtained from all participants as approved by each site’s institutional review board, and participants were financially compensated for their time.

Participants in the AURORA Study were enrolled from September 2017 to December 2020. A total of 2943 individuals were enrolled and a subset (n = 504) were invited to complete diffusion MRI at 2-weeks and/or 6-months after trauma. The present investigation on childhood adversity required participants to have childhood adversity data and structural neuroimaging data at both 2-weeks and 6-months post-trauma. Childhood adversity data consisted of either completion of a modified Childhood Trauma Questionnaire (mCTQ) or endorsement of traumatic exposure during childhood on the Life Events Checklist for DSM-5 (cLEC-5). Structural neuroimaging data consisted of diffusion weighted imaging (DWI) data that met the standards of quality control assessment (see below for details). In total, 114 participants had childhood adversity data and usable 2-week and 6-month neuroimaging data.

### Demographics

Demographic characteristics such as age, race, ethnicity, sex assigned at birth, income, and education were collected either in the ED or 2-week follow-up visit (Table 1).

**Table 1.** Demographic and ED trauma characteristics.

| **Characteristic** | **Mean (SD)**<br>**or n (%)** |
| --- | --- |
| **Sex at Birth** |  |
| Female | 80 (70.2) |
| Male | 34 (29.8) |
| **Age** | 37.7 (13.8) |
| **Race/Ethnicity** |  |
| Hispanic | 17 (14.9) |
| Non-Hispanic Black | 53 (46.5) |
| Non-Hispanic White | 38 (33.3) |
| Non-Hispanic Other | 5 (4.4) |
| Did not report | 1 (0.9) |
| **Education Level** |  |
| Some High School / Below | 5 (4.4) |
| High School / GED | 31 (27.2) |
| Some College | 42 (36.8) |
| Associate Degree | 13 (11.4) |
| Bachelor's Degree | 15 (13.2) |
| Graduate Degree | 8 (7.0) |
| **Annual Family Income** |  |
| Less than \$19,000 | 23 (20.2) |
| \$19,001 - \$35,000 | 37 (32.5) |
| \$35,001 - \$50,000 | 21 (18.4) |
| \$50,001 - \$75,000 | 10 (8.8) |
| \$75,001 - \$100,000 | 5 (4.4) |
| Greater than \$100,000 | 12 (10.5) |
| Did not report | 6 (5.3) |
| **Mechanisms of Injury** |  |
| Motor Vehicle Collision | 88 (77.2) |
| Non-motorized Collision | 2 (1.8) |
| Physical Assault | 9 (7.9) |
| Sexual Assault | 2 (1.8) |
| Fall < 10 feet / unknown | 6 (5.3) |
| Animal-related | 3 (2.6) |
| Burns | 1 (0.9) |
| Other | 3 (2.6) |

*ED* Emergency Department.

### Childhood adversity

Childhood adversity was assessed by two self-report questionnaires: a modified version of the Childhood Trauma Questionnaire (mCTQ) and endorsement of traumatic events on the Life Events Checklist for DSM-5 during childhood (cLEC-5). ***Modified CTQ.*** The AURORA study used the mCTQ (Bernstein et al., 2003) to assess exposure to maltreatment during childhood. The mCTQ included 11 items that covered 5 subtypes of maltreatment, including physical abuse, sexual abuse, emotional abuse, physical neglect, and emotional neglect. Participants self-reported on how often they experienced each type of maltreatment during childhood using a 5-point Likert-scale (0: never, 1: rarely, 2: sometimes, 3: often, 4: very often) within approximately two weeks after admission to ED. Items related to emotional neglect (i.e., “There was someone in your family who helped you feel that you were important or special”, “You felt loved”) and physical neglect (i.e., “You knew there was someone to take care of you and protect you”, “There was someone to take you to the doctor if you needed it”) were reverse coded so that greater scores represented higher levels of neglect. The total possible score ranged from 0 to 44 and the summed score was used in the present analyses. ***cLEC-5***. Childhood adversity was also assessed by endorsement of childhood trauma from Life Events Checklist for DSM-5 (cLEC-5) collected 8-weeks after ED admission. The LEC-5 is an established instrument for assessing exposure to a wide range of traumatic events (Weathers et al., 2013a). In the present study, we only considered events that happened during childhood. For each of the 17 traumatic events included in the survey (e.g., natural disaster, transportation accident, physical assault etc.), participants reported on whether they experienced it personally, whether they witnessed it happen to someone else, whether they learned about it happening to someone close to them, and whether they were exposed to details about it as part of their job (1: yes, 0: no). Participants also reported the age at which the first time the specific event occurred if any. Childhood adversity was calculated as the total number of traumatic events experienced at any of the four levels mentioned above between ages 3-10 to approximate the childhood developmental period. The total possible score ranged from 0 to 68. The summed raw score was used in the analyses.

Results of the two measures were included in the analyses separately given each encompasses potentially unique dimensions of childhood adversity despite some overlap (e.g., sexual assault). Specifically, cLEC-5 focused on direct or indirect exposure to traumatic events ranging from natural disasters to witnessing the death of a loved one. mCTQ, on the other hand, emphasized maltreatment experiences that may not necessarily reach a threshold of “trauma” within the DSM-5. The present study used both measures to maximize our ability to detect potentially unique aspects of childhood adversity on white matter microstructure.

### Posttraumatic Outcomes

Participants completed the Posttraumatic Stress Disorder checklist for DSM-5 (PCL-5; Blevins et al., 2015; Weathers et al., 2013b) to assess PTSD symptoms at 2-weeks, 6-months, and 12-months after the qualifying traumatic event. The PCL-5 included 20 items, and participants reported on how much they were troubled by the thoughts and feelings related to the traumatic event either in the past 2 weeks at 2-weeks post-trauma, or in the past 30 days at the following timepoints. The assessment used a Likert-scale of 4 (0: not at all, 1: a little, 2: some, 3: a lot, 4: extremely) and the total score (ranging from 0 to 80) was summed and used in the analyses. We used the calculated symptom difference score between 2-weeks and 6-months, and the symptom score at 12-months to assess the longitudinal posttraumatic outcomes. Descriptive statistics of the self-report measures are provided in Table 2.

**Table 2.**
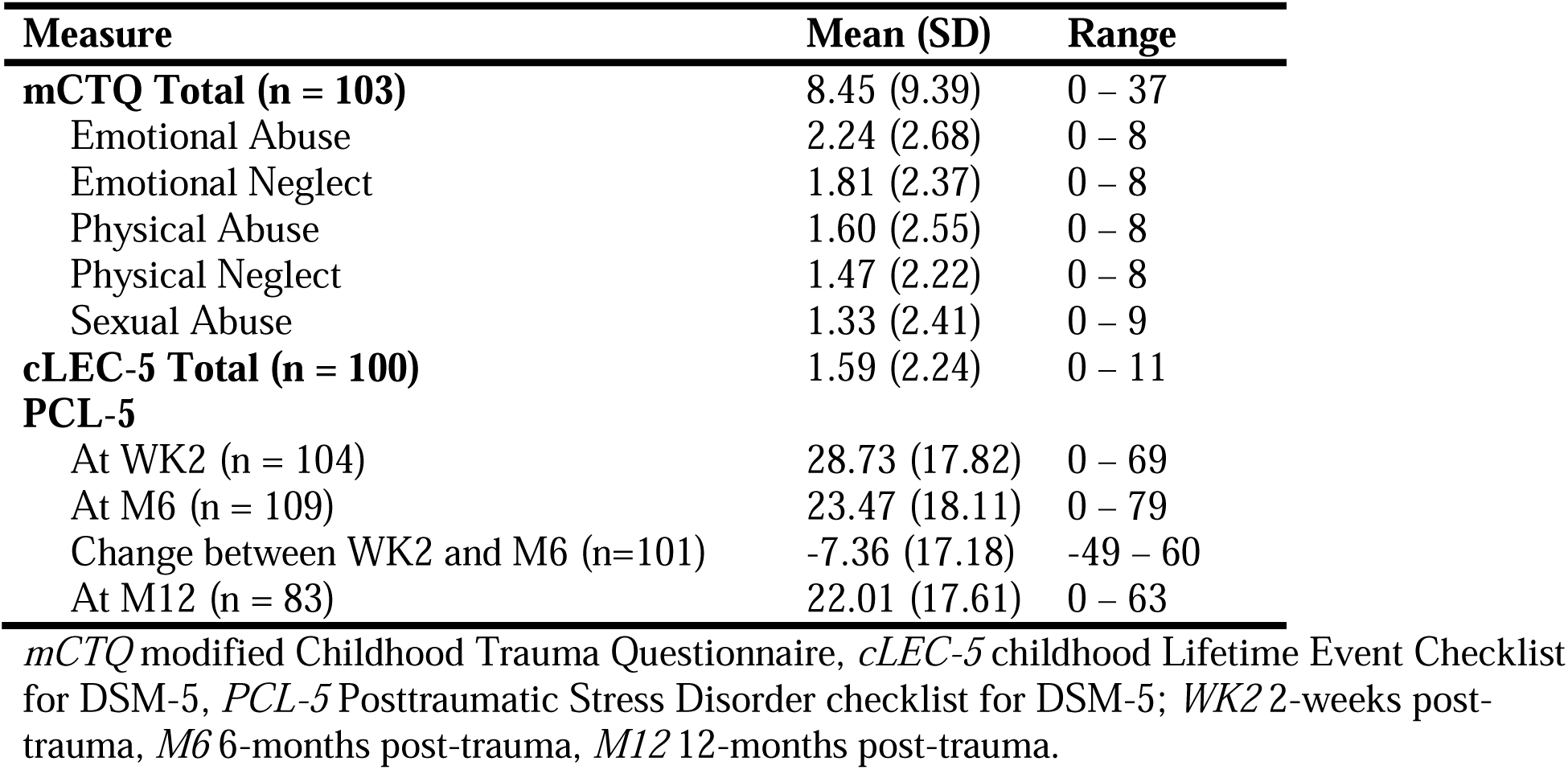
Distribution of childhood adversity scores.

### Diffusion weighted imaging

Diffusion weighted imaging (DWI) was conducted to obtain measures of white matter microstructure across five sites as previously reported (Wong et al., 2023). Data processing was completed following the recommendations of the ENIGMA consortium (http://enigma.ini.usc.edu/protocols/dti-protocols/). For data quality control, we first visually inspected diffusion weighted images and then calculated metrics of temporal signal-to-noise ratio (TSNR) and outlier maximum voxel intensity. We removed participants who demonstrated both: (a) TSNR values lower than 4.88 and (b) maximum voxel intensities greater than 5000 (Roalf et al., 2016). Motion and eddy current effects were reduced using the ‘eddy’ subroutine in the FMRIB Software Library (FSL; Smith et al., 2004, Andersson and Sotiropoulos, 2016). We also corrected susceptibility effects using nonlinear warping of the DWI data to the anatomical scans (Wang et al., 2017).

Data were then processed and analyzed using two different methods: region-of-interest (ROI) analysis with tract-based spatial statistics (TBSS) and whole-brain correlational tractography in DSI Studio. TBSS processing was conducted following the ENIGMA-DTI working group processing standards to extract FA values (Smith et al., 2006; Jahanshad et al., 2013). Briefly, all FA images were nonlinearly registered to the FA template in the Montreal Neurological Institute (MNI) space and averaged to generate a mean FA skeleton (Jahanshad et al., 2013). Each participant’s FA map was then projected onto the skeleton to extract FA values of ROIs based on the John’s Hopkins University (JHU) White Matter Atlas (Hua et al., 2008). The *a priori* ROIs for the present analysis included inferior fronto-occipital fasciculus (IFOF), sagittal stratum (SS), posterior thalamic radiation (PTR), uncinate fasciculus (UNC), cingulum-cingulate gyrus (CGC), and cingulum-hippocampus (CGH).

We further performed Q-space diffeomorphic reconstruction (QSDR; sampling length ratio = 1.25 mm, output resolution = 2 mm) for the 2-week and 6-month data, and generated a longitudinal QA database for whole-brain analyses (Yeh and Tseng, 2011; Yeh et al., 2010; Yeh et al., 2016). We completed longitudinal correlational tractography in DSI Studio (version “CHEN” October 2, 2023) to test whether quantitative anisotropy (QA) changes across any white matter tracts were associated with 1) childhood adversity or 2) changes in PTSD symptom between 2-weeks and 6-months following trauma. QA is an index that quantifies anisotropic diffusion along the primary fiber orientation in white matter tracts (Yeh et al., 2010). While both QA and FA measure water diffusion in white matter, FA is a normalized metric that reflects the proportion of anisotropic diffusion within a voxel, whereas QA removes isotropic diffusion and scales with spin density (Yeh et al., 2010; Yeh et al., 2013). This distinction makes QA less susceptible to partial volume effects, leading to improved performance in tractography (Yeh et al., 2013). Therefore, the present study used QA to assess white matter microstructure in the whole-brain tractography analyses.

### Statistical Analysis

We completed both ROI and whole-brain analyses. Observations whose mCTQ or cLEC score was more than 3 standard deviations away from the mean were identified as outliers and removed from analyses. We excluded 1 outlier in the analyses with mCTQ and 4 outliers with cLEC-5. We also excluded 2 outliers of PCL-5 difference score in the whole-brain correlational tractography analysis with PCL-5 changes. The ROI-based analyses were completed with R (R Core Team, 2023) and R Studio (Posit Team, 2023). We conducted linear regression models to test the effects of childhood adversity on longitudinal changes between 2-weeks and 6-months post-trauma (i.e., 6-months minus 2-weeks) in the bilateral tracts of interest extracted from the JHU atlas. Age, sex assigned at birth, and scanning site were included as covariates. In total, the two measures of childhood adversity - mCTQ and cLEC - each had 12 models generated. We further conducted FDR correction based on the Benjamini–Hochberg procedure to adjust for multiple comparisons (Benjamini & Hochberg, 1995).

The whole-brain longitudinal tractography analyses were completed using nonparametric Spearman partial correlation, a built-in statistical method in DSI Studio. We tested the correlation between childhood adversity or PCL-5 score difference over 2-weeks and 6-months and longitudinal change in QA values, and the effect of age, sex assigned at birth, and scanning site was removed by multiple regression. A deterministic fiber tracking algorithm was used to identify the tract associations above a T-score threshold of 2.5 (Yeh et al., 2013). A seeding region was placed at whole brain. Next, the tractography was refined by filtering the tracks with 16 iterations of topology-informed pruning (Yeh et al., 2019). Finally, a total of 4000 randomized permutations were applied to obtain the null distribution of the track length, and the false discovery rate (FDR) was estimated. Tracts that passed an FDR < 0.05 were considered significant.

In exploratory follow-up analyses to better understand our whole-brain effects, we selected tracts within the bilateral cingulum bundle or IFOF from tracts showing significant positive associations between mCTQ and QA increases. We then generated QA connectometry databases for the 2-week and 6-month data using the same settings as were used for the longitudinal connectometry database and extracted the average 2-week and 6-month QA values of the selected significant tracts of interest (i.e., cingulum bundle and IFOF). Multiple regression analyses were completed to examine the association between mCTQ and QA at 2-weeks and 6-months post-trauma of these tracts, covarying for age, sex assigned at birth, and scanning site. We further completed an additional whole-brain analysis similar to the above to investigate associations between changes in white matter microstructure and PTSD symptoms at 12-months post-trauma.

## Results

### Participant characteristics

Participant demographics are presented in Table 1. Table 2 presents the mean and standard deviation of the childhood adversity scores and PTSD symptom scores in the sample. Correlational analyses (Table S1) indicated that in the current sample, mCTQ was significantly positively correlated with PTSD symptom scores at 2-weeks and 6-months, but not at 12-months or with changes between 2-weeks and 6-months. cLEC-5 was not significantly correlated with PTSD symptom scores at any timepoints.

### ROI-based Analysis

We first tested whether childhood adversity was significantly associated with longitudinal changes in FA of the 12 bilateral ROIs (Table 3). Greater mCTQ was associated with decreases in FA of left IFOF, left PTR, left UNC, and left CGH (nominal p < 0.05). Similarly, greater cLEC-5 was significantly associated with decreases in FA of right UNC and left CGH (nominal p < 0.05). However, the observed effects in the analyses did not survive FDR correction.

**Table 3.** Effects of childhood adversity on FA changes in tracts of interest.

| | $\beta$ | t-statistic | *p*-values | FDR adjusted *p*-values |
| --- | --- | --- | --- | --- |
| **Models with mCTQ** |  |  |  |  |
| IFOF-L | -0.24 | -2.32 | 0.022* | 0.128 |
| IFOF-R | 0.00 | 0.00 | 0.996 | 0.996 |
| SS-L | -0.20 | -1.96 | 0.053 | 0.128 |
| SS-R | -0.07 | -0.65 | 0.515 | 0.618 |
| PTR-L | -0.23 | -2.27 | 0.025* | 0.128 |
| PTR-R | 0.02 | 0.24 | 0.812 | 0.886 |
| UNC-L | -0.20 | -2.06 | 0.042* | 0.128 |
| UNC-R | -0.09 | -0.84 | 0.401 | 0.601 |
| CGC-L | -0.11 | -1.14 | 0.259 | 0.444 |
| CGC-R | -0.16 | -1.55 | 0.124 | 0.248 |
| CGH-L | -0.20 | -2.04 | 0.044* | 0.128 |
| CGH-R | 0.07 | 0.72 | 0.474 | 0.618 |
| **Models with cLEC-5** |  |  |  |  |
| IFOF-L | -0.17 | -1.67 | 0.097 | 0.305 |
| IFOF-R | -0.06 | -0.56 | 0.574 | 0.677 |
| SS-L | -0.07 | -0.70 | 0.488 | 0.651 |
| SS-R | -0.12 | -1.21 | 0.228 | 0.421 |
| PTR-L | -0.11 | -1.09 | 0.281 | 0.421 |
| PTR-R | -0.16 | -1.65 | 0.102 | 0.305 |
| UNC-L | -0.13 | -1.35 | 0.179 | 0.421 |
| UNC-R | -0.22 | -2.23 | 0.028* | 0.170 |
| CGC-L | 0.05 | 0.50 | 0.621 | 0.677 |
| CGC-R | -0.12 | -1.14 | 0.258 | 0.421 |
| CGH-L | -0.22 | -2.32 | 0.022* | 0.170 |
| CGH-R | -0.02 | -0.15 | 0.881 | 0.881 |

FDR corrected *p*-values were calculated using the Benjamini-Hochberg method. \*Significant before FDR correction ( $p < 0.05$ ).
-*L* left tract, -*R* right tract; *IFOF* Inferior Fronto-Occipital Fasciculus, *SS* Sagittal Stratum, *PTR* Posterior Thalamic Radiation, *UNC* Uncinate Fasciculus, *CGC* Cingulum (Cingulate Gyrus), *CGH* Cingulum (Hippocampus).

### Whole-brain Analysis

#### Childhood Adversity and QA Change

We next completed whole-brain tractography analyses to investigate if changes in white matter QA between the 2-week and 6-month scans were associated with prior childhood adversity. Greater mCTQ was associated with increases in QA over time (i.e., greater at 6-months compared to 2-weeks) within the bilateral IFOF, left ILF, and bilateral cingulum bundles (Figure 1). We further observed that greater mCTQ was associated with decreases in QA over time (i.e., greater at 2-weeks compared to 6-months) within the dorsal cingulum (Figure 1). Additionally, greater cLEC-5 was associated with increases in QA over time within the bilateral IFOF, optic radiation (OR), left PTR, and bilateral cingulum bundles (Figure 1). We did not observe any associations where greater cLEC-5 was associated with decreased QA.

**Figure 1.**
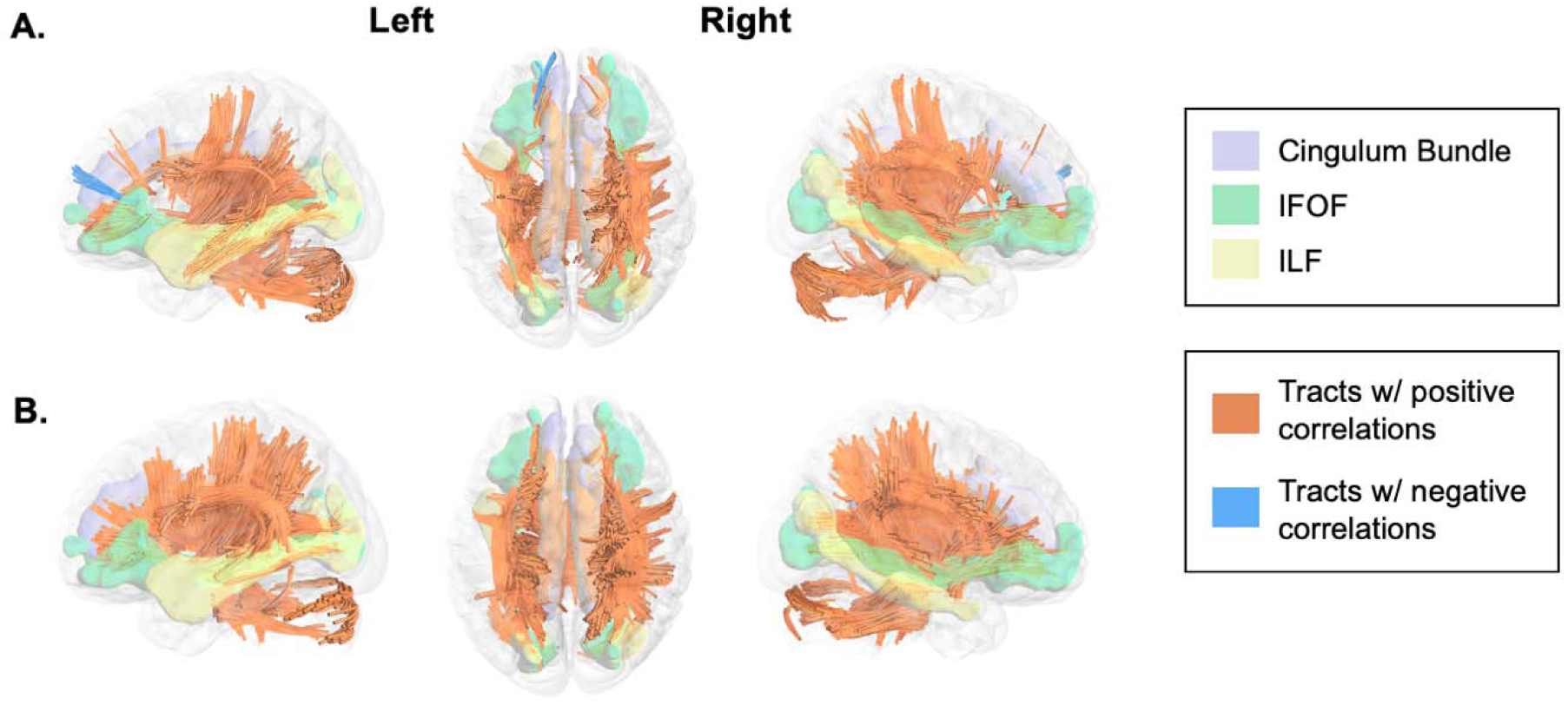
White matter changes after trauma are moderated by childhood adversity. Longitudinal correlational tractography analyses were completed to investigate the association between mCTQ (A) and cLEC-5 (B) scores with changes in quantitative anisotropy (QA). Across both measures, we observed significant positive associations between changes in QA between 2-weeks and 6-months and childhood adversity (orange tracts), and a negative association for mCTQ (blue tracts).

We then conducted exploratory follow-up analyses to test if the observed significant positive associations between mCTQ and QA increases over time were driven by QA values at 2-week or 6-month timepoint. Post hoc analyses revealed there were no significant associations between mCTQ and QA of tracts within bilateral cingulum bundle or IFOF at 2-weeks post-trauma (Table 4). However, mCTQ was significantly positively associated with QA of the bilateral cingulum bundle and right IFOF at 6-months (Table 4).

**Table 4.**
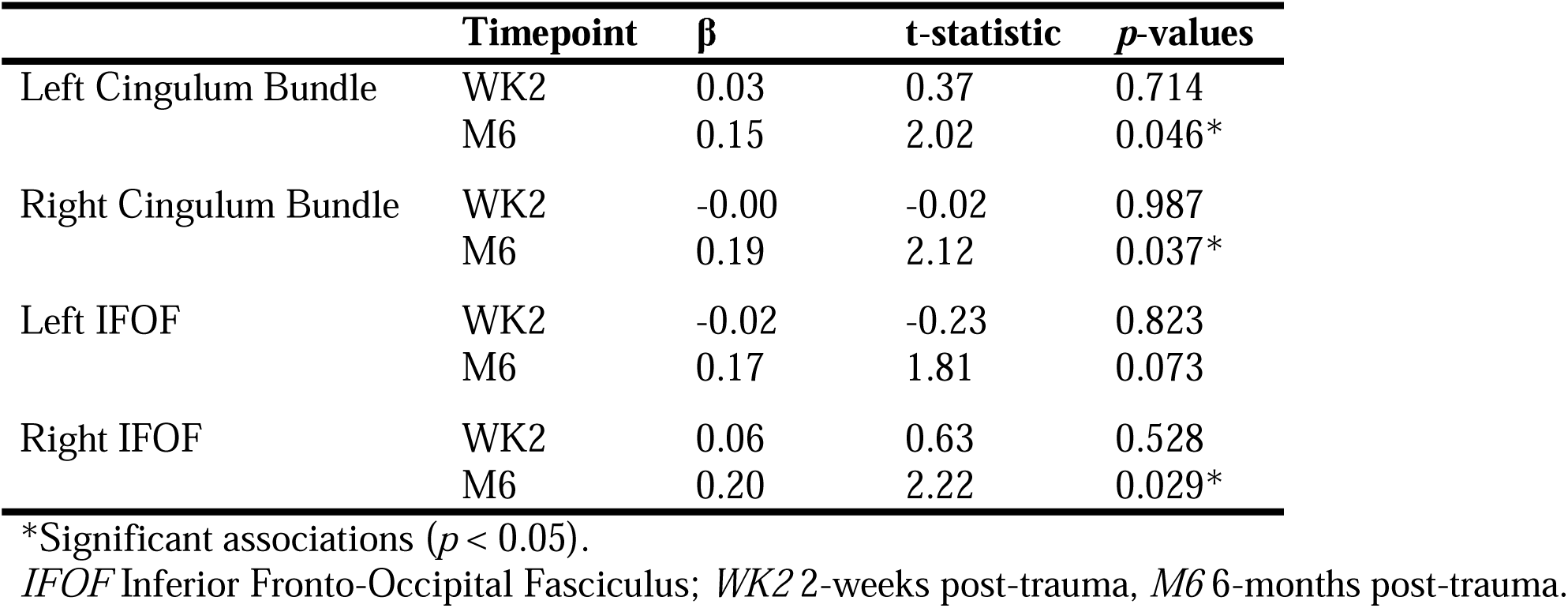
Associations between mCTQ and QA of significant tracts of interest at 2-weeks and 6-months post-trauma.

| | Timepoint | $\beta$ | t-statistic | p-values |
| --- | --- | --- | --- | --- |
| Left Cingulum Bundle | WK2 | 0.03 | 0.37 | 0.714 |
|  | M6 | 0.15 | 2.02 | 0.046* |
| Right Cingulum Bundle | WK2 | -0.00 | -0.02 | 0.987 |
|  | M6 | 0.19 | 2.12 | 0.037* |
| Left IFOF | WK2 | -0.02 | -0.23 | 0.823 |
|  | M6 | 0.17 | 1.81 | 0.073 |
| Right IFOF | WK2 | 0.06 | 0.63 | 0.528 |
|  | M6 | 0.20 | 2.22 | 0.029* |

\*Significant associations ( $p < 0.05$ ).
*IFOF* Inferior Fronto-Occipital Fasciculus; *WK2* 2-weeks post-trauma, *M6* 6-months post-trauma.

#### PTSD Symptom Change and QA Change

Next, we investigated if changes in white matter QA were associated with changes in PTSD symptoms. Whole-brain longitudinal correlational tractography with the PCL-5 change score (6-months minus 2-weeks) revealed that increases in PCL-5 scores were associated with increases in QA over time within bilateral cerebellum and forceps minor (Figure 2). We also found that increases in PCL-5 scores over time were associated with decreases in QA within forceps major, forceps minor, bilateral IFOF, cingulum bundle, ILF, middle longitudinal fasciculus, and left cerebellum (Figure 2).

**Figure 2.**
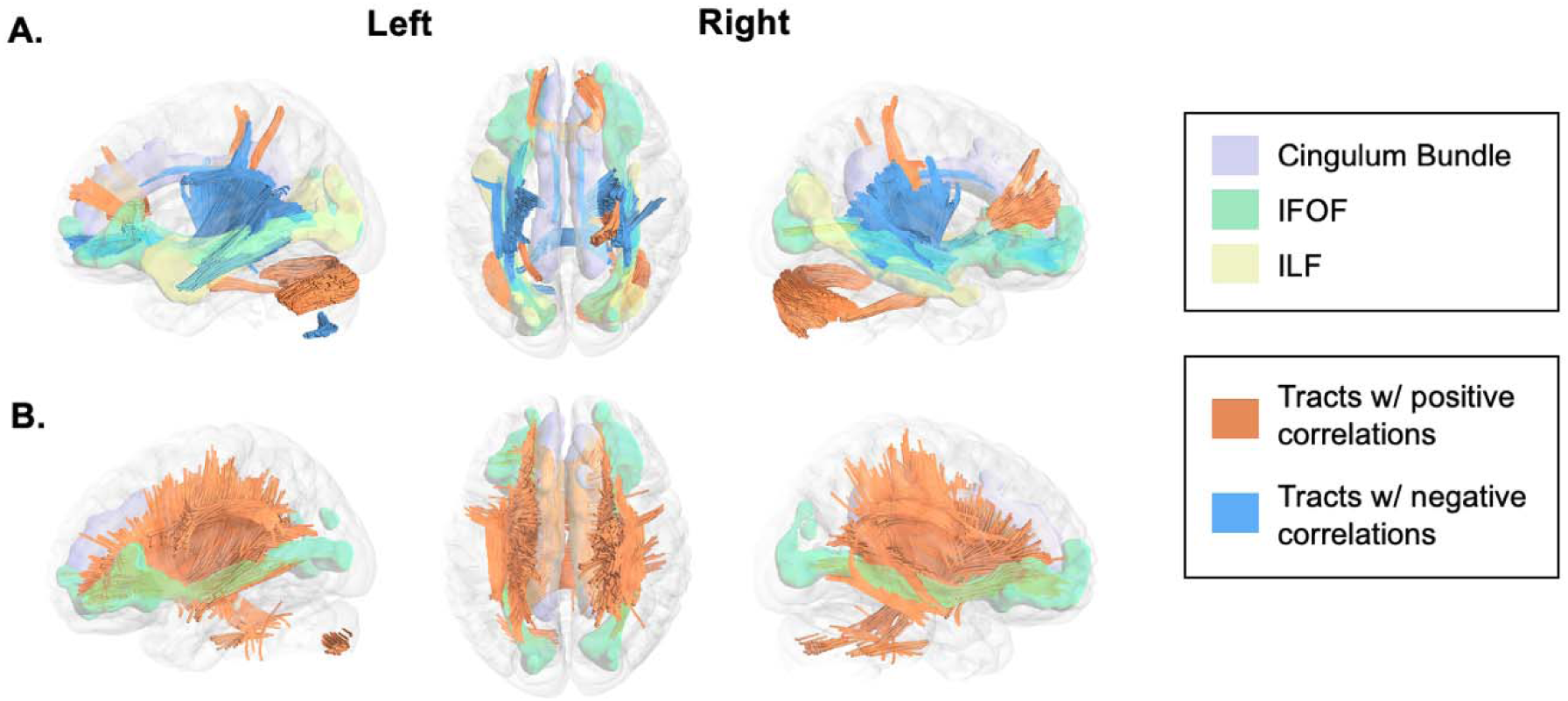
White matter changes after trauma are associated with post-trauma PTSD symptomatology. Longitudinal correlational tractography analyses were completed to investigate the association between changes in quantitative anisotropy (QA) and PCL-5 score changes between 2-weeks and 6-months (A) and PCL-5 scores at 12-months (B). We observed both significant positive (orange tracts) and negative (blue tracts) associations between changes in QA and changes in PCL-5 scores. Only positive associations (orange tracts) were observed with 12-month PCL-5 scores.

#### 12-Months PTSD Symptom and QA Change

Finally, we investigated if changes in white matter QA were predictive of future (i.e., 12-month) PTSD symptoms. Greater longitudinal increases in QA within the bilateral cingulum bundles, bilateral IFOF, right middle longitudinal fasciculus, corpus callosum, and SLF were associated with higher PCL-5 score at 12-months post-trauma (Figure 2). No negative associations were found.

## Discussion

Childhood adversity is associated with adulthood responses to traumatic stress, and recent evidence suggests that the association may be partially driven by variability in neural structure. However, limited research to date has investigated whether longitudinal changes in white matter microstructure, in the early months after trauma, may be related to prior exposure to childhood adversity. Understanding the potential associations of childhood adversity with changes in white matter may shed light on the neural mechanisms influencing susceptibility to post-traumatic stress. The present study investigated the associations between childhood adversity, changes in white matter microstructure, and PTSD symptoms in recent trauma survivors. Individuals with higher levels of childhood adversity showed increased microstructural integrity (indexed via QA) between 2-weeks and 6-months post-trauma within bilateral IFOF, OR, cingulum bundle, left ILF, and left PTR, and decreased QA within the dorsal cingulum. We further observed that increases in PTSD symptoms between 2-weeks and 6-months, as well as PTSD symptoms at 12-months, were mainly associated with increases in QA between 2-weeks and 6-months across several tracts, including bilateral cingulum bundle, IFOF, and ILF. The present findings demonstrate that childhood adversity is associated with white matter changes following adulthood trauma exposure, which – in turn – are associated with posttraumatic stress outcomes.

Our whole-brain tractography analysis revealed that the severity of childhood adversity, using either measures of childhood maltreatment or childhood trauma, was associated with changes in white matter QA across several white matter tracts. Specifically, greater adversity was associated with increases in QA during the months after trauma within bilateral IFOF, left ILF, and cingulum bundle as well as the optic and posterior thalamic radiations. The findings seem surprising within the context of prior cross-sectional studies that observed a negative association between childhood adversity and white matter integrity. A meta-analysis suggests individuals with maltreatment experiences in childhood, compared with non-maltreated controls, show reduced microstructure within bilateral fornix, OR, ILF, and IFOF (Lim et al., 2020). Our own prior work in the AURORA sample found childhood maltreatment load was negatively associated with 2-week post-trauma white matter microstructure in adulthood trauma survivors (Wong et al., 2023). The inconsistent findings may be partially attributable to the different timeframes. While the previous findings are mainly cross-sectional or specific to acute measure of white matter at one timepoint following adulthood trauma (i.e., 2-weeks post-trauma), the present study examined the prospective post-trauma changes in white matter microstructures in relation to childhood adversity. The present results are thus reflective of how childhood adversity may affect the neural responses to a later adulthood stress exposure, which may be different from the cross-sectional differences in white matter observed in individuals with exposure to childhood adversity. Another difference between the current report and prior research is that previous work commonly utilized diffusion tensor imaging metrics (e.g., FA) while the present findings were seen in QA. FA and QA summarize microstructural properties of white matter tracts using distinct methods and have differing biophysical mechanisms (Yeh et al., 2013) and thus the directionality of the present effects with QA may be different from prior reports focused on FA. However, prior work observed negative associations between QA and measures of adversity in a young adult sample including neighborhood disadvantage, income, and violence exposure (Bell et al., 2021). It is thus possible the present finding of increased QA in those with greater childhood adversity in the 6-months after an adulthood trauma exposure is driven by other factors than the microstructural index used here.

One potential explanation for the increases in QA is that adverse experiences in childhood may potentiate increases in white matter microstructural integrity in the late recovery period after adulthood traumatic stress. Childhood adversity is associated with adaptive neurobehavioral processes, such as heightened attentional bias towards threat-related visual stimuli (Pollak & Tolley-Schell, 2003; Shackman & Pollak, 2014). The neurobehavioral adaptions may offer protection against some future stressors while being maladaptive in other circumstances (Teicher et al., 2016). The IFOF and cingulum bundle are key tracts interconnecting visual and emotional processing regions (Forkel et al., 2014; Bubb et al., 2018) and–in the present study– microstructures of these tracts were associated with reports of childhood adversity. Further, we also observed that the association between childhood adversity and white matter QA changes in these tracts were primarily driven by the 6-month post-trauma assessment. Speculatively, it may be that individuals with prior adversity engage adaptive processes that could strengthen the fiber tracts by potentially triggering experience-driven myelination (Fields, 2015; Mount & Monje, 2017), in turn driving increased QA over time.

The possibility that childhood adversity may contribute to longer-term adaptive changes in white matter is partially supported by our finding that increased QA in tracts including cingulum, IFOF, and ILF was associated with decreased PTSD symptoms between 2-weeks and 6-months post-trauma. Childhood adversity (Lim et al., 2020; Huang et al., 2012; Hanson et al., 2015), trauma exposure (Hu et al., 2016; Wong et al., 2023; Harnett et al., 2020a; Li et al., 2016), and PTSD (Olson et al., 2017; Siehl et al., 2020; Ju et al., 2020; Dennis et al. 2021) are all associated with variability in tracts that support visual and emotional information processing (Harnett et al., 2024). However, there is limited research on how changes in white matter are associated with changes in PTSD symptoms over time. Treatment studies suggest greater FA reductions over time in the UNC, fornix, and stria terminalis are related to greater improvement in PTSD symptoms after treatment (Korem et al., 2024; Kennis et al., 2015). Further, increased cingulum FA over time is associated with resistance to PTSD treatment (Kennis et al., 2015). In the present study, individuals with greater increases in QA–previously associated with childhood adversity– showed greater reductions in PTSD symptoms. Our findings may suggest a partially protective effect of early adversity exposure on changes in PTSD symptoms over time. However, it should be noted that individuals with higher levels of adversity also exhibited greater PTSD symptoms at both 2-weeks and 6-months post-trauma. Thus, plasticity of white matter tracts may aid some individuals in preventing worsening of symptoms over time despite the potentiating effects of adversity on symptoms more generally. Interestingly, although increased white matter QA was associated with decreases in PTSD symptoms over 2-weeks and 6-months, increased QA was also associated with greater PTSD symptoms at 12-months. Taken together, the present findings suggest there are adaptive white matter changes facilitated by earlier stress exposure that help to mitigate increasing PTSD symptoms in the posttraumatic phase. However, such changes may–for some individuals–contribute to greater symptom expressions in the long term. Importantly, the present findings suggest there is temporal variability in white matter microstructure within the first year after trauma related to posttraumatic outcomes.

The present findings should be interpreted in the context of several limitations. First, there was limited information collected from participants on the timing of childhood adversity in our analyses. The timing of adverse experiences relative to development stages may have varying effects on white matter microstructure and the subsequent risk of developing PTSD in adulthood not considered in the present analysis. Prior research has highlighted sensitive periods in neurodevelopment when adverse experiences are particularly impactful on the development of tracts supporting threat response and sensorimotor functions (Gabard-Durnam & McLaughlin, 2019; Sisk et al., 2023). Second, while the present study is somewhat unique in its longitudinal design, neuroimaging was completed at only two time points over six months, which may not fully capture the potential dynamic structural changes associated with trauma exposure. Prior research suggests white matter exhibits dynamic and experience-dependent plasticity even in adulthood (Sampaio-Baptista & Johansen-Berg, 2017). Consequently, the microstructure of white matter may exhibit varying trajectories of changes following a traumatic event. Future studies would benefit from collecting diffusion weighted data at multiple timepoints, both in the early and later aftermath of trauma, to better elucidate the temporal patterns of white matter changes and their implications for posttraumatic stress symptoms. Multiple timepoints of data would also allow for sophisticated longitudinal models to take account of potential statistical limitations such as increased measurement error and omitted time invariant variables.

In conclusion, childhood adversity moderated the microstructural changes in white matter tracts implicated in threat and visual processing between 2-weeks and 6-months post-trauma, which were in turn related to fluctuations in, and persistence of, PTSD symptoms. These findings underscore the importance of investigating changes in white matter integrity over time in recent trauma survivors as it may serve as a possible neural pathway linking childhood adversity to elevated susceptibility to adulthood PTSD. Although more research is necessary to better characterize the temporal properties of white matter tracts, the present findings provide critical insights into the interconnection between childhood adversity, temporal variability in threat- and visual-related tracts, and posttraumatic stress responses in adulthood.

## Data Availability

Data used in this manuscript is available through the National Institute of Mental Health (NIMH) Data Archive (NDA). The NDA Collection for the AURORA Project can be found here: https://nda.nih.gov/edit_collection.html?id=2526. This content is solely the responsibility of the authors and may not reflect the official view of any of the funders or the Submitters submitting original data to NDA.

## Acknowledgements

The investigators wish to thank the trauma survivors participating in the AURORA Study. Their time and effort during a challenging period of their lives make our efforts to improve recovery for future trauma survivors possible. The present research was supported by a Brain Behavior Research Foundation Young Investigator Award (NGH) and the National Institute of Mental Health K01MH129828 (NGH). The AURORA study was supported by NIMH under U01MH110925, the US Army MRMC, One Mind, and The Mayday Fund. The DISENTANGLE study is a continuation of AURORA’s work and is supported by the United States Army Medical Research Acquisition Activity (USAMRAA) under Contract No. W81XWH22C012. Data used in this manuscript is available through the National Institute of Mental Health (NIMH) Data Archive (NDA). The NDA Collection for the AURORA Project can be found here: https://nda.nih.gov/edit_collection.html?id=2526. This content is solely the responsibility of the authors and may not reflect the official view of any of the funders or the Submitters submitting original data to NDA.

## Disclosures

- Dr. Huibregtse receives fellowship support from the National Institute of Mental Health (F32 MH134528).
- Dr. van Rooij is supported by the NIMH (K01MH121653).
- Dr. Lauren Lebois reports unpaid membership on the Scientific Committee for the International Society for the Study of Trauma and Dissociation (ISSTD), grant support from the National Institute of Mental Health (K01 MH118467), the Julia Kasparian Fund for Neuroscience Research, and the Trauma Scholars Fund. Dr. Lebois also reports spousal intellectual property payments from Vanderbilt University for technology licensed to Acadia Pharmaceuticals and spousal private equity in Violet Therapeutics, unrelated to the present work. Neither ISSTD nor NIMH was involved in the analysis or preparation of this manuscript.
- Dr. Jovanovic receives support from the National Institute of Mental Health, R01 MH129495.
- Dr. Neylan has received research support from NIH, VA, and Rainwater Charitable Foundation, and consulting income from Otsuka Pharmaceuticals.
- In the last three years Dr. Clifford has received research funding from the NSF, NIH and LifeBell AI, and unrestricted donations from AliveCor Inc, Amazon Research, the Center for Discovery, the Gates Foundation, Google, the Gordon and Betty Moore Foundation, MathWorks, Microsoft Research, Nextsense Inc, One Mind Foundation, and the Rett Research Foundation. Dr Clifford has financial interest in AliveCor Inc and Nextsense Inc. He also is the CTO of MindChild Medical with significant stock. These relationships are unconnected to the current work.
- Dr. Rauch reported serving as secretary of the Society of Biological Psychiatry; serving as a board member of Community Psychiatry and Mindpath Health; serving as a board member of National Association of Behavioral Healthcare; serving as secretary and a board member for the Anxiety and Depression Association of America; serving as a board member of the National Network of Depression Centers; receiving royalties from Oxford University Press, American Psychiatric Publishing Inc, and Springer Publishing; and receiving personal fees from the Society of Biological Psychiatry, Community Psychiatry and Mindpath Health, and National Association of Behavioral Healthcare outside the submitted work.
- Dr. Jones has no competing interests related to this work, though he has been an investigator on studies funded by AstraZeneca, Vapotherm, Abbott, and Ophirex.
- Dr. Pascual is president elect of the Society for Clinical Care Medicine.
- Dr. Datner serves as a Medical Advisor for Cayaba Care.
- In the past 3 years, Dr. Kessler was a consultant for Cambridge Health Alliance, Canandaigua VA Medical Center, Child Mind Institute, Holmusk, Massachusetts General Hospital, Partners Healthcare, Inc., RallyPoint Networks, Inc., Sage Therapeutics and University of North Carolina. He has stock options in Cerebral Inc., Mirah, PYM (Prepare Your Mind), Roga Sciences and Verisense Health.
- Dr. Koenen has done paid consulting for the US Department of Justice and Covington and Burling, LLP. She receives royalties from Oxford University Press and Guilford Press.
- Dr. Ressler has performed scientific consultation for Bioxcel, Bionomics, Acer, and Jazz Pharma; serves on Scientific Advisory Boards for Sage, Boehringer Ingelheim, Senseye, and the Brain Research Foundation, and he has received sponsored research support from Alto Neuroscience.
- Dr. McLean has served as a consultant for Walter Reed Army Institute for Research, Arbor Medical Innovations, and BioXcel Therapeutics, Inc.

## Notes

### Author Declarations

Written informed consent was obtained from all participants as approved by each sites institutional review board, with review completed by the overall study site, the Institutional Review Board at the University of North Carolina at Chapel Hill.

