## Supplemental Material for "Childhood adversity is associated with longitudinal white matter changes after adulthood trauma"

**Supplementary Information**

**Table S1. Correlations between childhood adversity and PCL-5.**

|  | **WK2** | |  | **M6** | |  | **Change** | |  | **M12** | |
| --- | --- | --- | --- | --- | --- | --- | --- | --- | --- | --- | --- |
|  | **r** | **p-values** |  | **r** | **p-values** |  | **r** | **p-values** |  | **r** | **p-values** |
| **mCTQ** | **0.32** | **0.001*** |  | **0.26** | **0.008*** |  | **-0.11** | **0.304** |  | **0.05** | **0.651** |
| **cLEC-5** | **0.17** | **0.102** |  | **0.03** | **0.743** |  | **-0.16** | **0.145** |  | **0.11** | **0.368** |

***mCTQ* modified Childhood Trauma Questionnaire, *cLEC-5* childhood Lifetime Event Checklist for DSM-5, *PCL-5* Posttraumatic Stress Disorder checklist for DSM-5; *WK2* PCL-5 score at 2-weeks, *M6* PCL-5 score at 6-months, *Change* PCL-5 score changes between 2-weeks and 6-months, *M12* PCL-5 score at 12-months; *significant correlations (p < .05).**
